# Patterns and predictors of COVID-19 vaccine uptake among United States active duty Service members, 2020-2022: Implications for future pandemics

**DOI:** 10.1101/2024.10.25.24316148

**Authors:** Erica Sercy, Laveta Stewart, Megan Clare Craig-Kuhn, Caryn Stern, Brock Graham, Amber Michel, Edward Parmelee, Simon Pollett, Timothy Burgess, David R Tribble

## Abstract

**Introduction:** Vaccine mandates have been used to minimize the duty days lost and deaths attributable to infectious disease among active duty Service members (ADSMs). In response to the global COVID-19 pandemic, in August 2021, the United States Department of Defense issued a COVID-19 vaccine mandate for all ADSMs. This study aimed to investigate COVID-19 vaccine uptake among the ADSM population, as well as factors associated with timing of COVID-19 vaccine receipt.

**Methods:** This study included ADSMs on active duty between 1/1/2020-6/30/2022. Univariate analyses investigated associations between demographic factors (age, sex, race, ethnicity, branch of service, rank, state of residence) and COVID-19 diagnosis with the following outcomes: 1) time to primary series initiation in relation to the DoD vaccine mandate, 2) time between doses of the two-dose primary series, and 3) time between booster eligibility and receipt

**Results:** A total of 1,799,466 ADSMs were included, with 90% receiving ≥1 COVID-19 vaccine dose during the study period and 77% initiating the primary series prior to the mandate. Over 80% of ADSMs received a complete primary series, with 96% of those adhering to the recommended regimen. History of COVID-19 diagnosis was associated with later receipt of all doses.

**Conclusions:** COVID-19 vaccine uptake was high among all ADSMs, with the majority initiating the primary series before the mandate. The high vaccine uptake among ADSMs shown here may be used as a guide to both military and civilian pandemic policy and outreach efforts related to enhanced vaccine uptake.

## Introduction

Historically, the United States (U.S.) military system has required specific vaccinations to aid in combat readiness and preserve health.^1,2^ During the 1918 influenza pandemic, ∼20-40% of U.S. Army and Navy personnel experienced symptomatic illness, associated with ∼8.7 million lost duty days and 26,000 deaths.^3^ In response, an influenza vaccination requirement was issued in 1945 for all military personnel, and similar requirements have continued to be issued to cover the vaccine schedule recommended by the U.S. Centers for Disease Control and Prevention (CDC) and in response to emerging infectious agents.^3,4^ Vaccines currently required by the U.S. Department of Defense (DoD) for all Service members prior to entry or at accession include adenovirus, hepatitis A, hepatitis B, influenza, measles/mumps/rubella (MMR), meningococcal, tetanus-diphtheria (Tdap), and varicella.^1,4,5^ Periodic administration of other vaccines is required in specific risk-based circumstances, including yellow fever, typhoid, Japanese encephalitis, and polio, as well as annual influenza vaccination.^6^ Prior to the COVID-19 pandemic, all vaccines required by the DoD had full U.S. Food and Drug Administration (FDA) approval.^4,5^

The first case of COVID-19 was reported in November 2019 in Wuhan, China, followed by rapid spread worldwide.^7^ Development of a COVID-19 vaccine began in late March 2020,^8^ leading to the first Emergency Use Authorization (EUA) by the FDA in mid-December 2020 for a two-dose vaccine series, with additional EUAs subsequently being granted to several other vaccine products.^9–11^ Between January 2020 and August 2021, there were ∼190,000 COVID-19 cases among active duty and Reserve/Guard Service members.^12^ By July 2021, even in the absence of FDA approval, a large percentage (∼67%) of the active duty population had received at least one dose of a COVID-19 vaccine.^13^ Following the first FDA approval of a COVID-19 vaccine on 23 August 2021 (Pfizer),^14^ the DoD issued a mandate on 24 August 2021 that all active duty Service members must be vaccinated against COVID-19 or face discharge to “protect the health and readiness of the force.”^15^ In September 2021, the first EUA for a booster dose was issued.^16^

Despite high levels of early uptake of the COVID-19 vaccine among active duty Service members, several challenges to full vaccination have presented. In surveys of military personnel, 20-40% reported COVID-19 vaccine hesitancy, noting concerns about “short-term side effects,” “long-term side effects,” “vaccine effectiveness,” “being infected with COVID-19 from the vaccine,” “worry about misinformation/political agenda,” and “lack of trust in the government.”^17–19^ In addition, because two of the three most commonly available vaccines in the U.S. in the early pandemic comprised a two-dose series (Pfizer, Moderna), attrition after the first dose led to incomplete vaccination: as of May 11, 2023, 81% of the general U.S. population had received at least one dose, but only 70% completed a two-dose primary series.^20^ To inform preparedness plans for future vaccine-preventable infectious threats, this study aimed to describe the patterns and predictors of COVID-19 vaccine receipt among active duty Service members in the Military Health System (MHS).

## Methods

### Study Population and Variables

Data without personal identifiers were from the MHS Data Repository (MDR), a claims-based centralized repository that includes healthcare data on all beneficiaries of the MHS, including active duty Service members, Reserve/Guard members, veterans/retirees, and dependents. This research received an exempt determination by the Uniformed Services University of the Health Sciences Human Research Protection Program.

This study included active duty Service members and Reserve/Guard members on active duty (collectively ADSMs) at any time in 1/1/2020-6/30/2022. Individuals were excluded if they had missing age or age <17 or >65. Only ADSMs who received all doses from products with EUAs in the U.S. (Pfizer, Moderna, Janssen/Johnson & Johnson) were included. Individuals were excluded if they met any of the following conditions that suggested documentation error: 1) a first recorded dose date prior to the EUA date (Pfizer: 12/11/2020; Moderna: 12/18/2020; Janssen/Johnson & Johnson: 2/27/2021); 2) booster dates prior to the first booster EUA (Pfizer: 9/22/2021); 3) <17 days between primary series doses (Pfizer, Moderna); or 4) <5 months (150 days, Pfizer, Moderna) or <2 months (60 days, Janssen/Johnson & Johnson) between completion of the primary series and the first booster (Supplementary Figure 1).

The following variables were collected on all individuals: demographics (age, sex, race, ethnicity, branch of service, rank, state of residence), all COVID-19 vaccine doses (date, manufacturer), and COVID-19 diagnosis (infection criteria met, diagnosis date). State of residence was defined using permanent residence for Reserve/Guard members and assigned unit location for individuals on active duty; residence was measured at COVID-19 diagnosis date for those with infection during the study period and most recent address (end of study period or end of active duty) for individuals with no COVID-19 diagnosis during the study period. COVID-19 diagnosis was defined as either lab confirmed, i.e., documented positive PCR or antigen test, or using ICD-10-CM codes (Supplementary Table 1). The index date of infection was the first date meeting either lab or ICD-10-CM criteria; for individuals with multiple COVID-19 diagnoses, only the earliest diagnosis date was considered.

A high degree of collinearity was detected between state of residence and multiple other variables (race, branch of service, rank) in the dataset. It is likely that much of the collinearity between state of residence and branch of service is attributable to the differential presence of military bases in each state (Supplementary Figure 2). Because previous studies have reported wide geographic variation in COVID-19 waves, variants, and vaccination rates across the U.S.,^21–23^ it was determined that state of residence should be retained as a variable in the final analyses. Therefore, to account for collinearity, findings are reported overall and stratified by state of residence. Only results for select states with the largest ADSM populations are displayed in the stratified analyses.

### Statistical Analyses

Associations of demographics and COVID-19 diagnosis with the following outcomes were examined: 1) initiation of the primary series in relation to the DoD COVID-19 vaccine mandate, 2) completion of a two-dose primary series in the manufacturer-recommended timeframe, and 3) days between booster eligibility and receipt. Timing of primary series initiation was defined as receipt of a first COVID-19 vaccine primary series dose prior to compared to after the DoD mandate. Timing of primary series completion was defined as occurring within the period recommended by the vaccine manufacturer (Pfizer or Moderna; ≤56 days between the first and second dose) compared to late (>56 days). An individual’s booster eligibility date was defined as the later of the following: 1) 22 September 2021 (the EUA for the first booster) or 2) the completion date of the primary series plus 5 months (Pfizer or Moderna primary series) or 2 months (Janssen/Johnson & Johnson primary dose). Univariate associations with each outcome were assessed in the population overall and stratified by state of residence using chi-square tests, Mann-Whitney U tests, and unadjusted logistic or generalized linear regression modeling. Adjusted regression analyses were not performed because of multicollinearity that remained in multivariable models after stratifying by state of residence. All statistical analyses were conducted using SAS Studio (SAS Institute, Inc., Cary, NC).

## Results

The study population included 1,799,466 ADSMs. Most of the population was age 17-49 (98%), male (82%), white (71%), non-Hispanic (83%), an enlisted-level Service member (83%), and in the Army (39%), Navy (23%), or Air Force (22%) (Supplementary Table 2). The seven states with the largest ADSM populations were California (12% of the full population), Texas (9%), Virginia (8%), North Carolina (7%), Georgia (5%), Florida (5%), and Washington (4%). In the full population, 23% of ADSMs received a complete primary COVID-19 vaccine series and at least one booster, 61% received a complete primary series with no booster, 5% received an incomplete primary series, and 10% had no record of receiving one of the three COVID-19 vaccine products evaluated here (Table 1). Florida (90%) and Virginia (90%) had the highest percentages of individuals who received a complete primary series, followed by Washington (86%), California (84%), North Carolina (83%), Texas (83%), and Georgia (82%) (P<0.0001).

**Table 1.** Vaccine status, overall and in select states of residence.

|  | All | California | Texas | Virginia | North Carolina | Georgia | Florida | Washington | P<sup>a</sup> |
| --- | --- | --- | --- | --- | --- | --- | --- | --- | --- |
|  | N=1,779,466 | N=213,128 | N=162,709 | N=146,939 | N=124,573 | N=84,380 | N=83,156 | N=76,683 |  |
| Vaccine history |  |  |  |  |  |  |  |  | <0.0001 |
| Complete primary series, ≥1 booster | 415,359<br>(23.3%) | 40,689<br>(19.1%) | 30,439<br>(18.7%) | 38,632<br>(26.3%) | 16,341<br>(13.1%) | 12,957<br>(15.4%) | 17,822<br>(21.4%) | 17,678<br>(23.1%) |  |
| Complete primary series, no booster | 1,082,882<br>(60.9%) | 139,186<br>(65.3%) | 103,903<br>(63.9%) | 93,209<br>(63.4%) | 87,201<br>(70.0%) | 55,906<br>(66.3%) | 57,223<br>(68.8%) | 48,164<br>(62.8%) |  |
| Incomplete primary series | 96,707<br>(5.4%) | 8,534<br>(4.0%) | 11,294<br>(6.9%) | 2,838<br>(1.9%) | 3,483<br>(2.8%) | 4,882<br>(5.8%) | 2,076<br>(2.5%) | 3,132<br>(4.1%) |  |
| No record of vaccination | 184,518<br>(10.4%) | 24,719<br>(11.6%) | 17,073<br>(10.5%) | 12,260<br>(8.3%) | 17,548<br>(14.1%) | 10,635<br>(12.6%) | 6,035<br>(7.3%) | 7,709<br>(10.1%) |  |
| ADSMs who initiated a primary series<sup>b</sup> | N=1,594,948 | N=188,409 | N=145,636 | N=134,679 | N=107,025 | N=73,745 | N=77,121 | N=68,974 |  |
| Primary series initiation timing<sup>c</sup> |  |  |  |  |  |  |  |  | <0.0001 |
| Pre-mandate | 1,230,623<br>(77.2%) | 147,271<br>(78.2%) | 108,818<br>(74.7%) | 108,256<br>(80.4%) | 75,147<br>(70.2%) | 55,405<br>(75.1%) | 56,734<br>(73.6%) | 58,248<br>(84.5%) |  |
| Post-mandate | 364,325<br>(22.8%) | 41,138<br>(21.8%) | 36,818<br>(25.3%) | 26,423<br>(19.6%) | 31,878<br>(29.8%) | 18,340<br>(24.9%) | 20,387<br>(26.4%) | 10,726<br>(15.6%) |  |
| ADSMs who completed a two-dose primary series<sup>d</sup> | N=1,444,775 | N=171,978 | N=131,004 | N=124,257 | N=98,234 | N=66,499 | N=70,350 | N=60,715 |  |
| Primary series completion timing<sup>e</sup> |  |  |  |  |  |  |  |  | <0.0001 |
| Recommended | 1,389,085<br>(96.2%) | 165,245<br>(96.1%) | 124,558<br>(95.1%) | 120,944<br>(97.3%) | 94,342<br>(96.0%) | 63,816<br>(96.0%) | 68,325<br>(97.1%) | 60,715<br>(97.0%) |  |
| Late | 55,690<br>(3.9%) | 6,733<br>(3.9%) | 6,446 (4.9%) | 3,313<br>(2.7%) | 3,892<br>(4.0%) | 2,683<br>(4.0%) | 2,025<br>(2.9%) | 1,862<br>(3.0%) |  |
| ADSMs who received ≥1 booster dose following primary series completion | N=415,359 | N=40,689 | N=30,439 | N=38,632 | N=16,341 | N=12,957 | N=17,822 | N=17,678 |  |
| Time from booster eligibility to receipt (days), median (IQR) | 90 (68-120) | 98 (71-125) | 85 (59-115) | 88 (63-118) | 94 (63-127) | 96 (70-125) | 95 (70-120) | 98 (72-121) | <0.0001 |

<sup>a</sup>P values are for comparisons across the seven states listed in the table. <sup>b</sup>Includes those who initiated either a one-dose or two-dose
primary series. <sup>c</sup>Pre-mandate is defined as receipt prior to 23 August 2021, post-mandate is defined as on or after 23 August 2021.
<sup>d</sup>Includes only those who completed a two-dose primary series. <sup>e</sup>Primary series completion in the recommended timing was defined as ≤56 days between the first and second dose; late is defined as >56 days between the first and second dose.
ADSMs, active duty Service members; IQR, interquartile range.

Among the 1,594,948 ADSMs who initiated a primary series, 77% initiated prior to the DoD mandate (Figure 1 and Table 1). Primary initiation timing differed by state of residence, with Washington (85%), Virginia (80%), and California (78%) having the highest percentages of residents initiating prior to the mandate. Individual-level factors significantly associated with initiation of the primary series prior to the mandate included older age (OR=2.46, 95% CI 2.38-2.54), female sex (OR=1.04, 95% CI 1.03-1.05), American Indian or Alaska Native (OR=1.06, 95% CI 1.02-1.10), Asian or Pacific Islander (OR=2.02, 95% CI 1.98-2.05), or other (OR=1.34, 95% CI 1.32-1.37) race compared to white, Hispanic ethnicity (OR=1.03, 95% CI 1.02-1.04), Coast Guard (OR=1.54, 95% CI 1.50-1.58) or Navy (OR=1.54, 95% CI 1.52-1.55) branch of service compared to Army, and officer (OR=4.28, 95% CI 4.22-4.35) or warrant officer (OR=2.03, 95% CI 1.96-2.10) rank compared to enlisted (Table 2 and Supplementary Table 3). Black or African American (OR=0.75, 95% CI 0.74-0.76) race, Air Force (OR=0.88, 95% CI 0.87-0.89) or Marine Corps (OR=0.58, 95% CI 0.57-0.59) branch of service, and COVID-19 diagnosis prior to initiating the primary series (OR=0.55, 95% CI 0.54-0.55 for lab confirmed, OR=0.51, 95% CI 0.50-0.52 for ICD-10-CM defined) were associated with lower odds of initiating before the mandate. The higher odds of pre-mandate initiation for older ADSMs, Asian or Pacific Islanders, and officer or warrant officers held across all states; likewise, the lower odds of pre-mandate initiation for blacks or African Americans, those in the Marine Corps, and those with a COVID-19 diagnosis prior to initiating the primary series held across all seven states (Table 2 and Supplementary Table 4).

**Figure 1.**
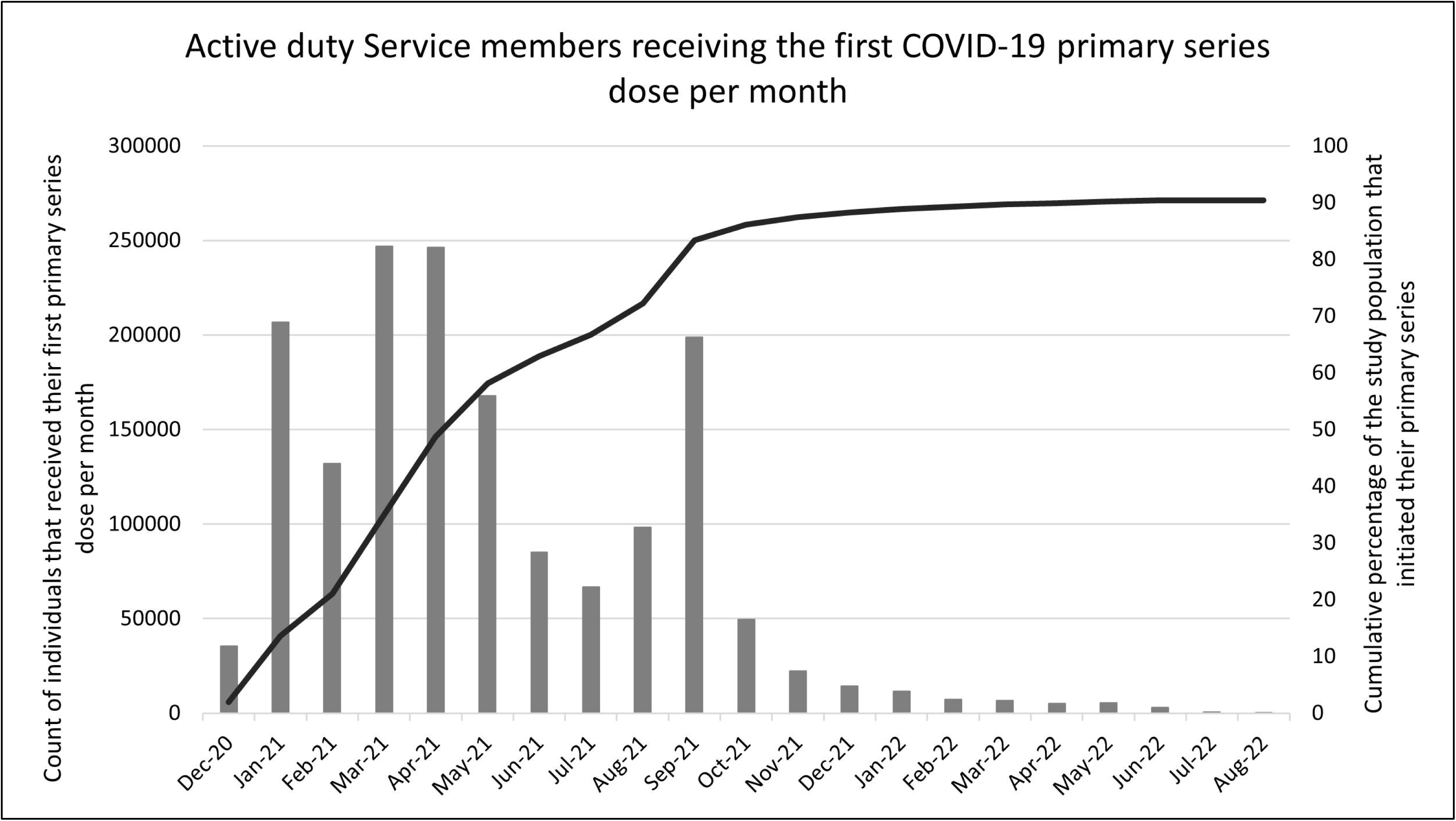
Monthly frequency of COVID-19 vaccine initiation

**Table 2.**
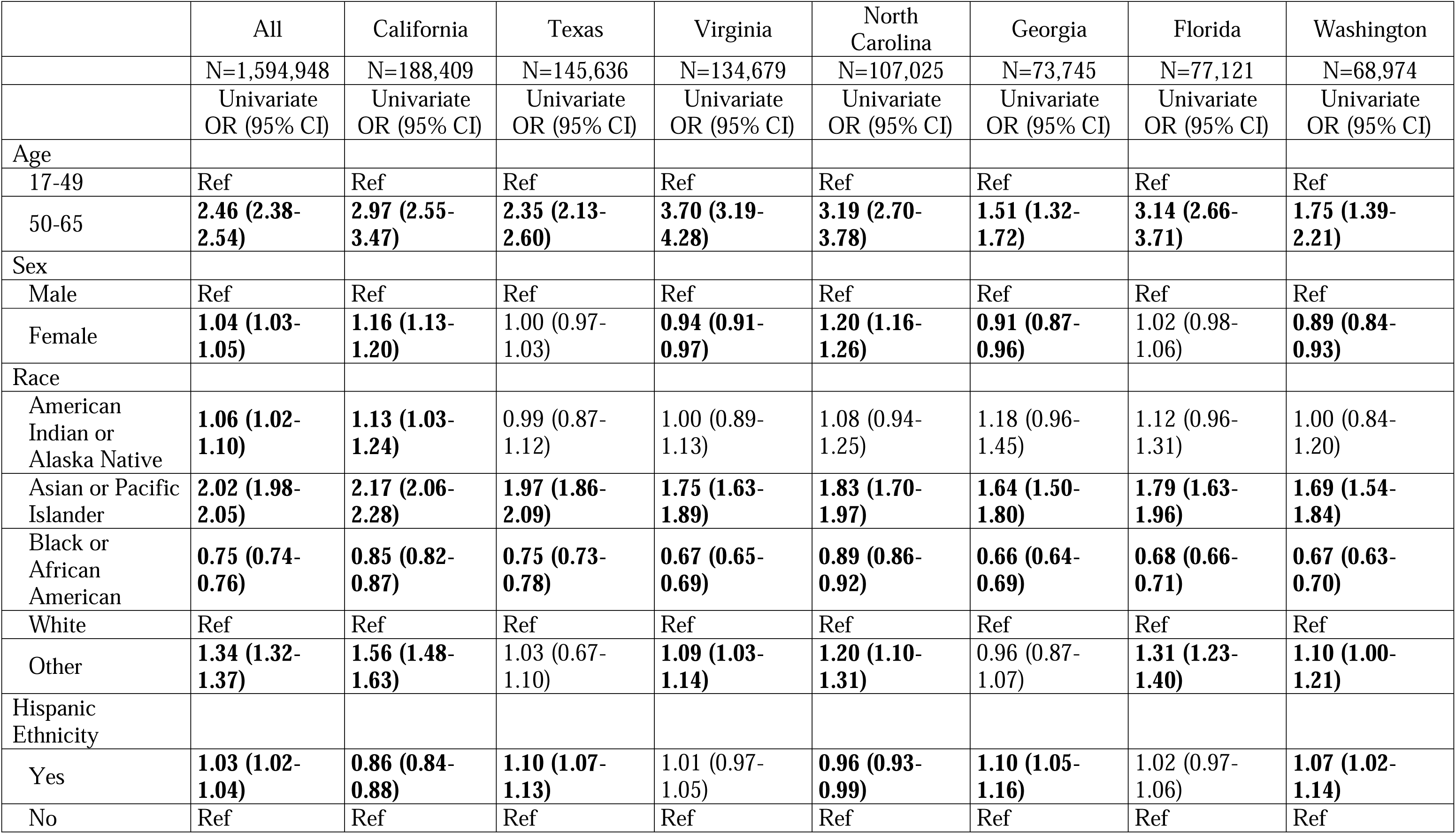
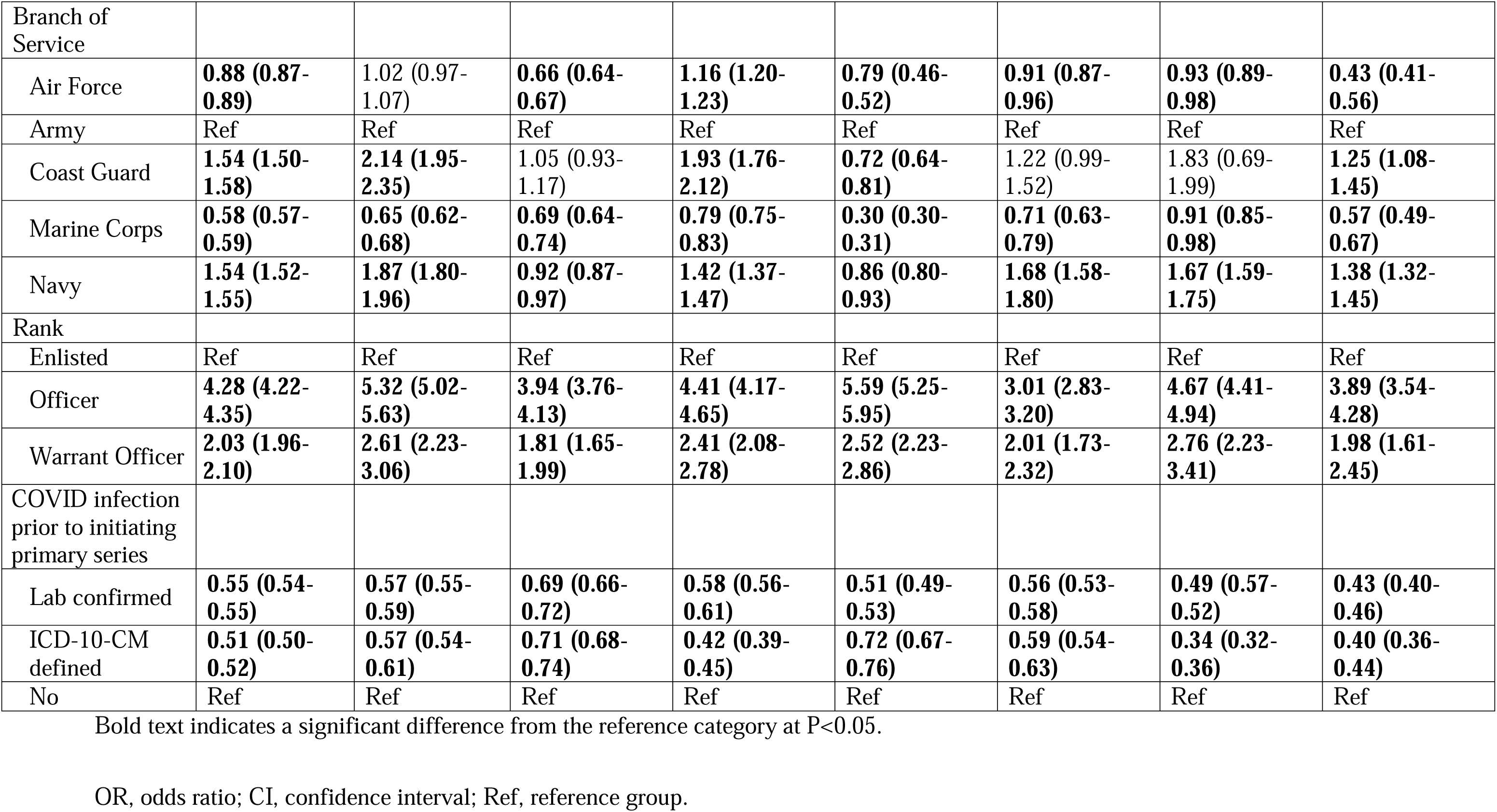
Unadjusted odds of initiating the primary vaccine series prior to the DoD mandate, overall and in select states of residence^a^.

Among those ADSMs who completed a two-dose primary series, 96% received the second dose within the recommended timeframe (Table 1). The percentage of ADSMs completing their primary series in the recommended time differed significantly by state, ranging from a low of 95% in Texas to a high of 97% in Virginia, Florida, and Washington. Female sex (OR=1.04, 95% CI 1.02-1.06), Asian or Pacific Islander (OR=1.19, 95% CI 1.15-1.24), and other (OR=1.25, 95% CI 1.20-1.30) race compared to white, Air Force (OR=1.33, 95% CI 1.30-1.36), Coast Guard (OR=1.07, 95% CI 1.02-1.13), Marine Corps (OR=1.17, 95% CI 1.14-1.20), and Navy (OR=1.80, 95% CI 1.76-1.85) branch of service compared to Army, and officer (OR=1.44, 95% CI 1.41-1.48) rank compared to enlisted had higher odds of completing the primary series in the recommended timeframe (Supplementary Tables 3 and 5). Black or African American race (OR=0.90, 95% CI 0.88-0.92) and a COVID-19 diagnosis between the first and second primary doses (lab confirmed, OR=0.23, 95% CI 0.21-0.24; ICD-10-CM defined, OR=0.23, 95% CI 0.22-0.25) were associated with lower odds of completing the second primary dose in the recommended time. The lower odds of primary series completion in the recommended time for COVID-19 diagnosis between primary doses held across all seven states (Supplementary Tables 5 and 6).

Among ADSMs who completed a primary series and received at least one booster, the median time between booster eligibility and receipt was 90 days (Table 1). This median time was significantly different across states, ranging from 85 days in Texas to 98 days in California and Washington. Individual factors associated with shorter time between eligibility and receipt included older age (the older age group got their booster a mean of 18.78, 95% CI 17.96-19.59, days before the younger group), female sex (5.93, 95% CI 5.53-6.33, days before males), other race (0.81, 95% CI 0.16-1.47, day before whites), Coast Guard (6.72, 95% CI 5.71-7.73, days before Army) branch of service, and officer (16.13, 95% CI 15.78-16.49, days before enlisted) or warrant officer (7.86, 95% CI 6.71-9.01, days before enlisted) rank (Table 3 and Supplementary Table 3). Factors associated with longer time between booster eligibility and receipt included American Indian or Alaska Native race (4.23, 95% CI 2.58-5.87, days after whites), Hispanic ethnicity (4.20, 95% CI 3.76-4.65, days after non-Hispanics), Air Force (0.69, 95% CI 0.28-1.10, days after Army), Marine Corps (18.21, 95% CI 17.51-18.91, days after Army) and Navy (7.74, 95% CI 7.33-8.16, days after Army) branch of service, and COVID-19 diagnosis between completion of the primary series and the booster (40.72, 95% CI 40.07-41.36, days after the no-infection group for lab confirmed; 22.49, 95% CI 21.61-23.37, days after the no-infection group for ICD-10-CM defined). The associations of older age and female sex with fewer days between booster eligibility and receipt and the associations of COVID-19 diagnosis between the primary series and booster with longer time between eligibility and receipt were consistent across all states (Table 3 and Supplementary Table 7).

**Table 3.**
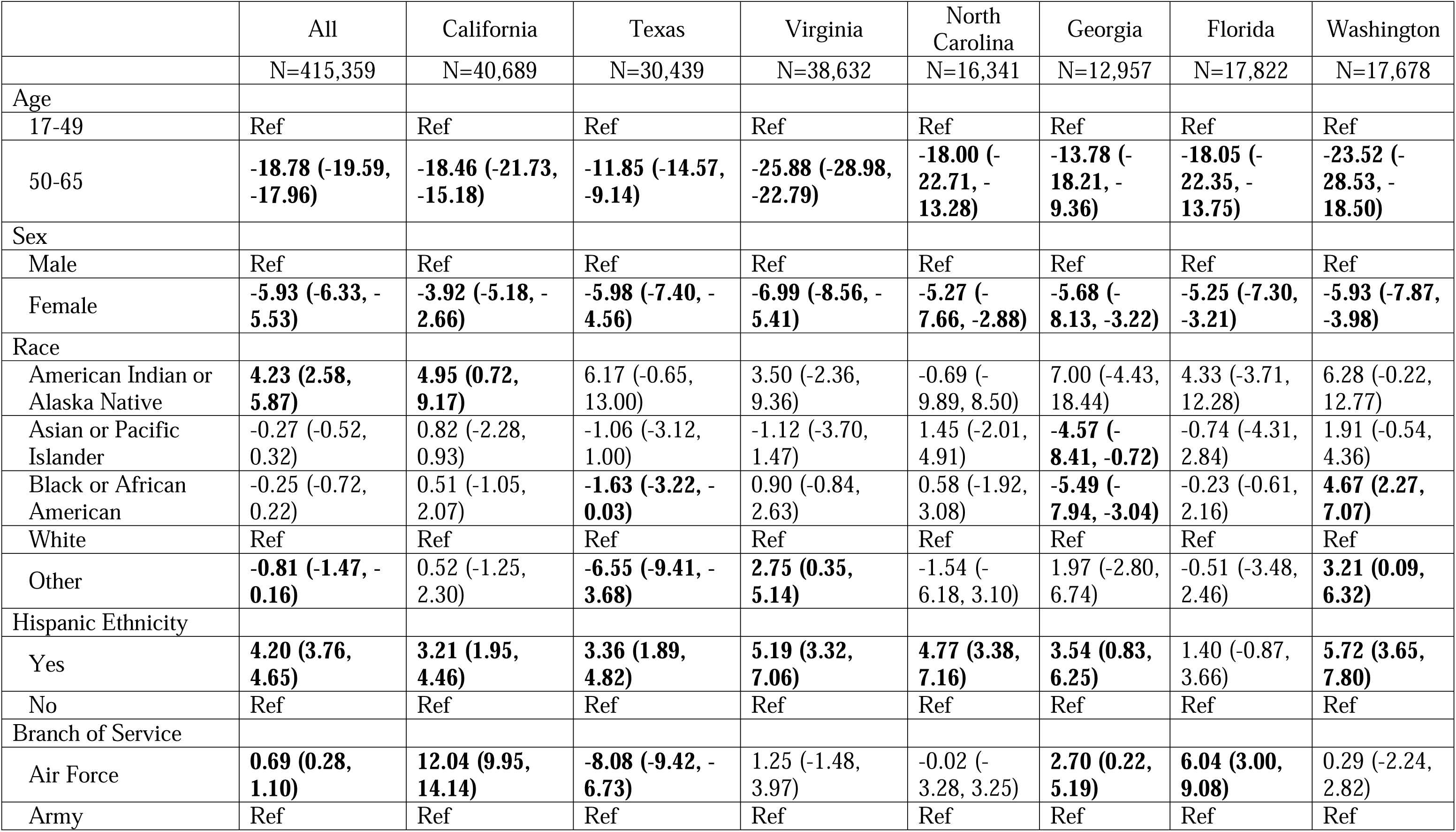
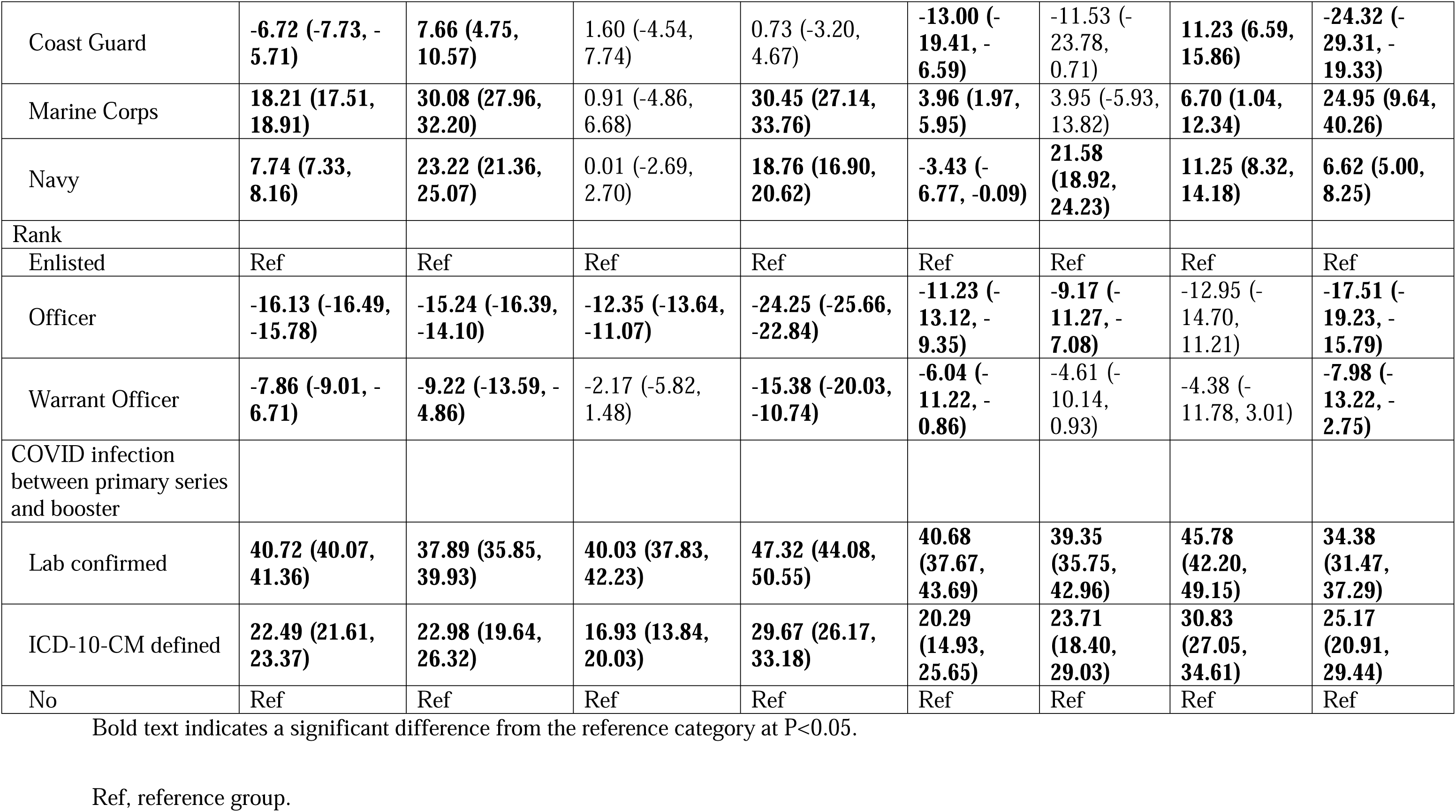
Unadjusted difference in the mean days between booster eligibility and receipt, overall and in select states of residence.

## Discussion

In this study, which aimed to evaluate U.S. military deployment of the COVID-19 vaccine during the pandemic, results showed that most of the ADSMs who initiated a COVID-19 primary series did so prior to the DoD vaccine mandate, at over three-fourths (77%) of the population reported here. The mandate resulted in a further substantial increase, with 87% of the population initiating by the end of December 2021 and 90% at the end of the study period (Figure 1). This is higher than in the overall adult U.S. population, where ∼78% of individuals had received at least one vaccine dose by the end of the study period (30 June 2022).^20^

Trends in vaccine uptake varied over time between the overall U.S. and ADSM populations. Both groups showed a large spike in primary series initiation soon after the EUAs in January-April 2021 (Figure 1).^20^ However, initiation among ADSMs thereafter showed a single notable spike after the DoD mandate in August 2021 (Figure 1). Trends in overall U.S. vaccine initiation appeared to be more responsive to COVID-19 variant waves,^24^ with later peaks in initiation corresponding to the Delta (June-November 2021) and early Omicron (December 2021-February 2022) waves in the U.S.^25^ The ADSM population did not show similar responses to these waves, although the DoD mandate was issued during the Delta wave in the U.S., and so the peak in initiation after the mandate may have been affected by both factors.

Some associations of demographic factors with vaccine timing outcomes were consistent across all states, such as the increased odds of higher-ranking ADSMs initiating their primary series before the mandate, completing their second dose in the recommended time, and receiving their booster closer to their eligibility date. Additionally, COVID-19 diagnosis during a relevant timeframe was consistently associated with less-favorable vaccine metrics. However, this delay is consistent with CDC recommendations, stating that individuals should not receive a COVID-19 vaccine during active COVID-19 disease and should delay their vaccine until after their isolation period is over: 7-10 days after symptom onset or first positive test and up to 20 days for those with severe illness.^26^ The CDC additionally suggested during the early pandemic that individuals consider delaying COVID-19 vaccination until 3 months after symptom onset or a first positive test, in an effort to preserve vaccine supply and capitalize on natural immunity from infection.^27^

Other associations varied by state, likely reflecting complex factors that interact to influence vaccination behavior, including differential vaccine messaging and deadlines within each branch of service, ^28^ overall vaccine messaging and environment by state,^29–31^ and requirements related to deployment,^32^ among many other secular factors (e.g., issues with vaccine supply shortages in certain regions).^33,34^

In a complementary manuscript, Dullea et al. (unpublished observations) described demographic factors in the ADSM population associated with completion of the COVID-19 primary vaccine series after the DoD mandate compared to those who remained unvaccinated in 12/11/2020-1/1/2022. Their findings showing an association between history of COVID-19 infection and unvaccinated status mirror the findings reported here. Further, Dullea et al. found that ADSMs who completed their series in response to the mandate were more likely to be black or African American, Hispanic, male, or of officer rank compared to those who remained unvaccinated. As our findings here reported that ADSMs who were black or African American, Hispanic, or male were less likely to initiate their primary series pre-mandate, the Dullea et al. manuscript shows that the DoD mandate “picked up” those individuals who did not initiate pre-mandate and reduced disparities in vaccine status across demographic factors by the end of the observation period, further emphasizing the effectiveness of the mandate among this population.

Limitations of COVID-19 vaccine data include highly variable vaccine availability and eligibility for the primary series and boosters over the course of the pandemic. For example, these analyses considered booster eligibility as the later of 22 September 2021 (the first booster EUA) or 5 months after primary series completion. However, on 22 September 2021, the Pfizer booster was recommended only for select high-risk individuals.^35^ Therefore, most of the young, healthy population included here was likely not recommended to receive a booster dose on this date; however, to standardize analyses, this EUA date was utilized as the booster eligibility date across the population. This likely resulted in overestimation of times between booster eligibility and receipt.

## Conclusions

Vaccine development during the COVID-19 pandemic proceeded at an extraordinarily rapid pace, with EUAs issued only ∼9 months after the official start of the pandemic. The DoD has historically required select vaccines among ADSMs, but these requirements have applied to vaccines with full FDA approval. The findings here showed that ADSMs were highly responsive to seeking COVID-19 vaccination during the EUA period, prior to FDA approval. The DoD COVID-19 vaccine mandate was subsequently successful at further raising the already high coverage among ADSMs. Overall, although uptake differed significantly by branch of service, rank, and state of residence, the data described here indicate an effective vaccine implementation effort by the DoD during the COVID-19 pandemic, leading to COVID-19 vaccine coverage that exceeded coverage in the civilian population, as well as earlier uptake than the overall U.S. population.

## Data Availability

All data produced in the present study are available upon reasonable request to the authors.

## Acknowledgements

Support for this work was provided by the Infectious Disease Clinical Research Program (IDCRP), a Department of Defense program executed through the Uniformed Services University of the Health Sciences, Department of Preventive Medicine and Biostatistics through a cooperative agreement with The Henry M. Jackson Foundation for the Advancement of Military Medicine, Inc. (HJF). This project has been funded by the National Institute of Allergy and Infectious Diseases, National Institutes of Health, under Inter-Agency Agreement Y1-AI-5072, and the Defense Health Program, U.S. DoD, under award HU0001190002.

