## Supplementary Tables and Figures for "Patterns and predictors of COVID-19 vaccine uptake among United States active duty Service members, 2020-2022: Implications for future pandemics"

Supplementary Figure 1. Study population flow chart

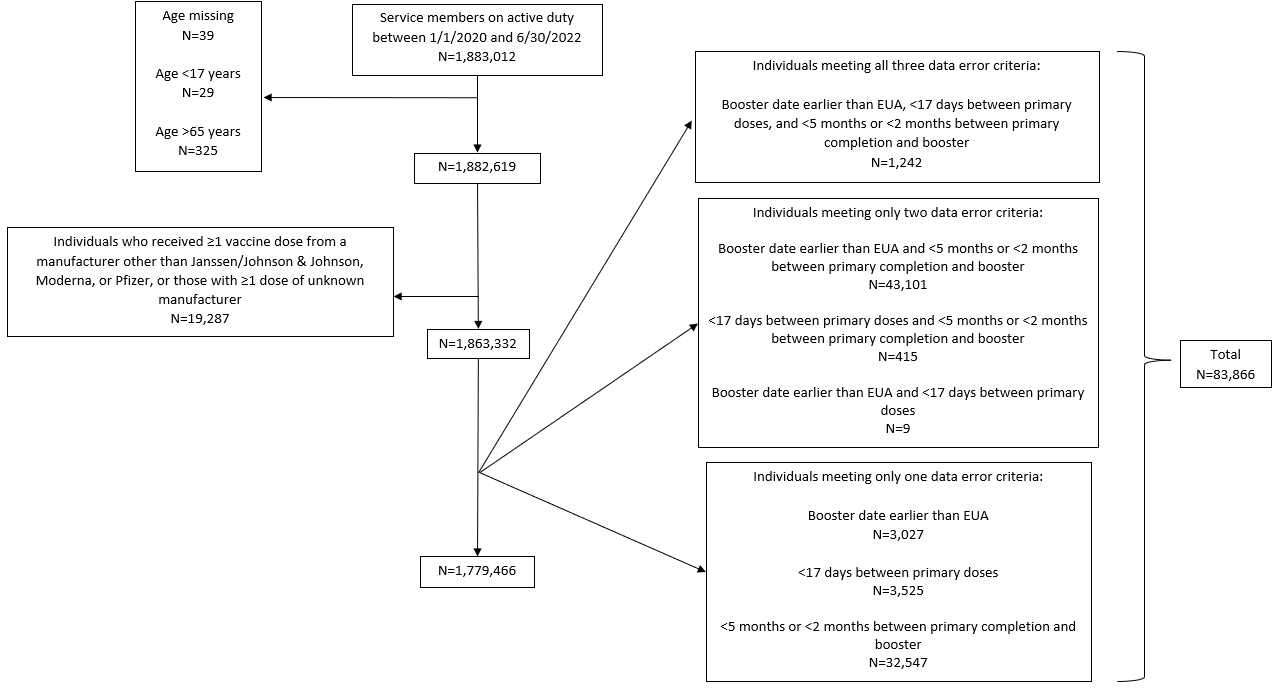

Supplementary Figure 2. Distribution of active duty Service member populations by branch in the study population

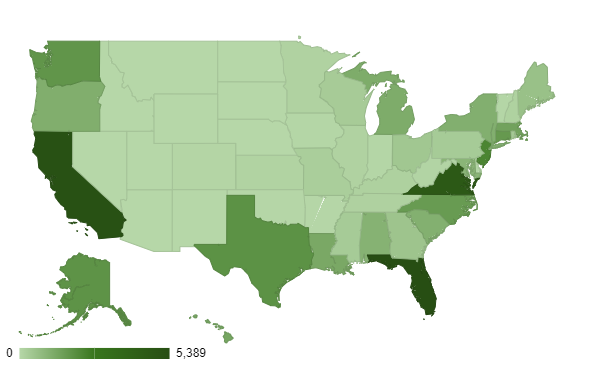

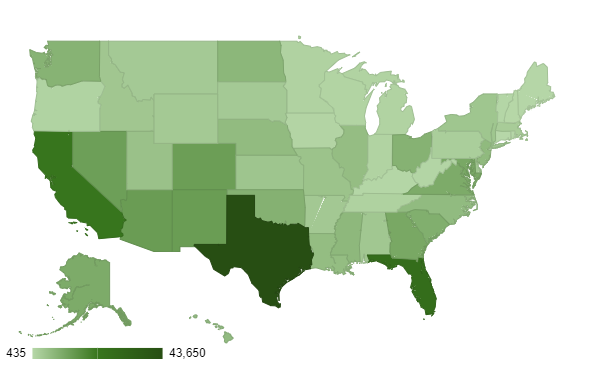
Air Force	Army	Coast Guard

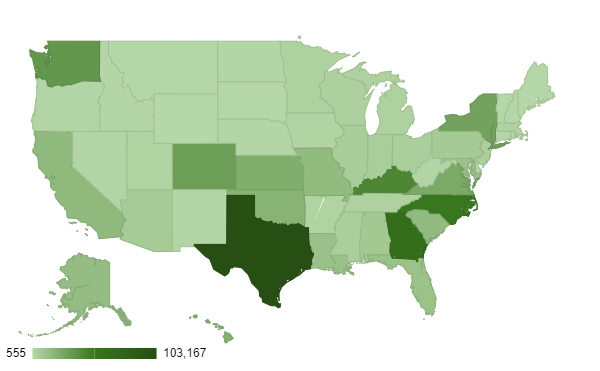

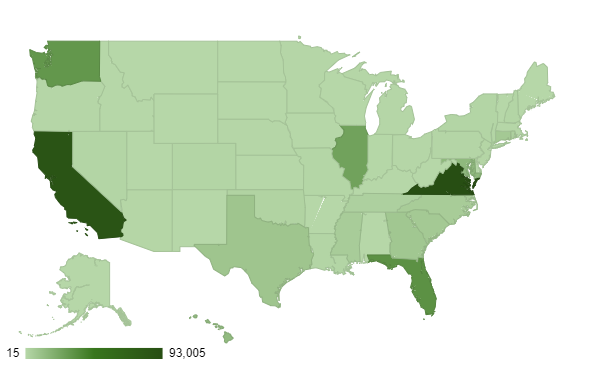

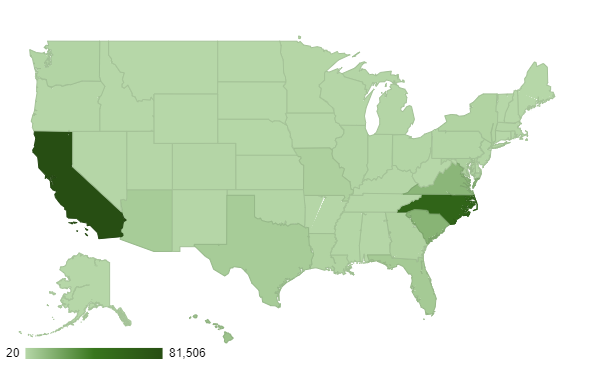
Marines	Navy

Supplementary Table 1. Identification and definition of individuals with COVID-19

| **ICD-10-CM Code** | **Definition** | **Requirements** |
| --- | --- | --- |
| Confirmed Cases | | |
| n/a | Positive lab results from molecular and/or antigen test. Positive tests are taken as cases even when negative tests are recorded within a short time frame. | - Earliest positive test in study period |
| Probable Cases | | |
| U07.1 | 2019-nCoV acute respiratory disease, COVID-19, virus identified | - Earliest occurrence of any one of these codes in study period |
| U07.2 | COVID-19, virus not identified (clinically diagnosed) (*this code is not used in the US, but is used internationally*) |  |
| J12.82 | Pneumonia due to coronavirus disease 2019 |  |
| J12.81 | Pneumonia due to SARS-associated coronavirus |  |
| B34.2 | Coronavirus, unspecified | - Earliest occurrence of either code in study period |
| B97.2 | Coronavirus as the cause of disease classified elsewhere |  |
| Possible Cases: Code Z20.822 and/or Z20.828 indicating contact with and (suspected) exposure to a SARS-CoV-2 infected person AND any **one** of the following conditions within ±14 days | | |
| J12.89 | Other viral pneumonia | - Earliest occurrence of any one of these codes in study period - Must be within ±14 days of suspected exposure |
| J12.9 | Viral pneumonia unspecified |  |
| J16.8 | Pneumonia due to other specified infectious organism |  |
| J17 | Pneumonia in diseases classified elsewhere |  |
| J18.0 | Bronchopneumonia, unspecified organism |  |
| J18.1 | Lobar pneumonia, unspecified organism |  |
| J18.2 | Hypostatic pneumonia, unspecified organism |  |
| J18.8 | Other pneumonia, unspecified organism |  |
| J18.9 | Pneumonia, unspecified organism |  |
| J84.111 | Idiopathic interstitial pneumonia not otherwise specified |  |
| J80 | Acute Respiratory Distress Syndrome |  |
| J06.0 | Acute laryngopharyngitis |  |
| J06.9 | Acute upper respiratory infection, unspecified |  |
| J20.8 | Acute bronchitis due to other specified organisms |  |
| J20.9 | Acute bronchitis, unspecified |  |
| J22 | Unspecified acute lower respiratory infection |  |
| J40 | Bronchitis, not specified as acute or chronic |  |
| J96.0 | Acute respiratory failure |  |
| J98.8 | Other specified respiratory disorders |  |
| M35.81 | Multisystem inflammatory syndrome |  |
| R06.03 | Acute Respiratory Distress |  |
| R05 | Cough |  |
| R06.02 | Shortness of Breath | - Earliest occurrence of any one of these codes in study period - Must be within ±14 days of suspected exposure - For each code in this group identified, there must be no record of the condition between Jan. 1, 2017 and Dec. 31, 2019 |
| R06.00 | Dyspnea, unspecified |  |
| R06.09 | Other forms of dyspnea |  |
| R43.0 | Anosmia |  |
| R43.2 | Ageusia |  |
| R43.8 | Other disturbances of smell and taste |  |
| R43.9 | Unspecified disturbances of smell and taste |  |
| R41.0 | Disorientation, unspecified |  |
| R41.82 | Altered mental status, unspecified |  |
| R41.9 | Unspecified symptoms and signs involving cognitive functions and awareness |  |
| R07.89 | Other chest pain |  |
| R07.9 | Chest pain, unspecified |  |
| Possible Cases: Code Z20.822 and/or Z20.828 indicating contact with and (suspected) exposure to a SARS-CoV-2 infected person AND any **two** of the following conditions, all within ±14 days of each other | | |
| R50.9 | Fever, unspecified | - Earliest occurrence of two of these codes within ±14 days of each other in study period - Must also be within ±14 days of suspected exposure |
| R68.83 | Chills (without fever) |  |
| M79.1 | Myalgia |  |
| R51 | Headache |  |
| R07.0 | Pain in throat |  |
| R11.0 | Nausea |  |
| R11.10 | Vomiting unspecified |  |
| R11.11 | Vomiting without nausea |  |
| R11.2 | Nausea with vomiting, unspecified |  |
| R19.7 | Diarrhea, unspecified |  |
| R53.83 | Other fatigue |  |
| R09.81 | Nasal congestion |  |

Supplementary Table 2. Demographics of the study population, overall and in select states of residence

|  | Full population | California | Texas | Virginia | North Carolina | Georgia | Florida | Washington |
| --- | --- | --- | --- | --- | --- | --- | --- | --- |
|  | N=1,779,466 | N=213,128 | N=162,709 | N=146,939 | N=124,573 | N=84,380 | N=83,156 | N=76,683 |
| Age |  |  |  |  |  |  |  |  |
| 17-49 | 1,739,941 (97.8%) | 210,865 (98.9%) | 158,996 (97.7%) | 143,668 (97.8%) | 123,031 (98.8%) | 82,647 (98.0%) | 81,505 (98.0%) | 75,752 (98.8%) |
| 50-65 | 39,525 (2.2%) | 2,263 (1.1%) | 3,713 (2.3%) | 3,271 (2.2%) | 1,542 (1.2%) | 1,733 (2.1%) | 1,651 (2.0%) | 931 (1.2%) |
| Sex |  |  |  |  |  |  |  |  |
| Male | 1,465,248 (82.3%) | 178,844 (83.9%) | 130,740 (80.4%) | 116,059 (79.0%) | 110,238 (88.5%) | 72,090 (85.4%) | 67,249 (80.9%) | 64,094 (83.6%) |
| Female | 314,210 (17.7%) | 34,284 (16.1%) | 31,969 (19.7%) | 30,880 (21.0%) | 14,335 (11.5%) | 12,290 (14.6%) | 15,910 (19.1%) | 12,589 (16.4%) |
| Race |  |  |  |  |  |  |  |  |
| American Indian or Alaska Native | 18,424 (1.1%) | 3,073 (1.5%) | 1,395 (0.9%) | 1,977 (1.4%) | 1,127 (0.9%) | 623 (0.8%) | 890 (1.1%) | 1,067 (1.4%) |
| Asian or Pacific Islander | 108,193 (6.2%) | 17,129 (8.1%) | 10,479 (6.5%) | 8,035 (5.5%) | 5,278 (4.3%) | 3,802 (4.6%) | 3,588 (4.4%) | 6,400 (8.4%) |
| Black or African American | 293,864 (16.8%) | 28,614 (13.5%) | 31,712 (19.8%) | 29,673 (20.4%) | 18,263 (14.9%) | 18,636 (22.3%) | 14,161 (17.3%) | 10,970 (14.4%) |
| White | 1,240,259 (70.7%) | 147,678 (69.7%) | 110,823 (69.2%) | 93,947 (64.6%) | 94,765 (77.3%) | 57,926 (69.4%) | 56,915 (69.6%) | 53,351 (69.9%) |
| Other | 94,019 (5.4%) | 15,333 (7.2%) | 5,855 (3.7%) | 11,762 (8.1%) | 3,137 (2.6%) | 2,489 (3.0%) | 6,283 (7.7%) | 4,515 (5.9%) |
| Hispanic Ethnicity |  |  |  |  |  |  |  |  |
| Yes | 307,225 (17.5%) | 53,107 (25.1%) | 35,448 (22.1%) | 22,374 (15.4%) | 21,474 (17.5%) | 11,462 (13.7%) | 14,594 (17.8%) | 12,916 (16.9%) |
| No | 1,449,578 (82.5%) | 158,813 (74.9%) | 125,087 (77.9%) | 123,125 (84.6%) | 101,252 (82.5%) | 72,184 (86.3%) | 67,319 (82.2%) | 63,463 (83.1%) |
| Branch of Service |  |  |  |  |  |  |  |  |
| Air Force | 395,819 (22.4%) | 22,806 (10.8%) | 43,650 (26.9%) | 10,215 (7.0%) | 6,724 (5.4%) | 10,932 (13.1%) | 26,694 (32.2%) | 8,475 (11.1%) |
| Army | 686,913 (38.9%) | 16,385 (7.7%) | 103,167 (63.7%) | 24,758 (16.9%) | 51,445 (41.4%) | 62,482 (74.7%) | 11,814 (14.3%) | 34,218 (44.7%) |
| Coast Guard | 45,217 (2.6%) | 5,219 (2.5%) | 1,917 (1.2%) | 4,642 (3.2%) | 1,660 (1.3%) | 515 (0.6%) | 5,389 (6.5%) | 1,822 (2.4%) |
| Marine Corps | 241,194 (13.6%) | 81,506 (38.5%) | 4,709 (2.9%) | 13,794 (9.4%) | 59,314 (47.7%) | 1,994 (2.4%) | 5,514 (6.7%) | 1,250 (1.6%) |
| Navy | 398,625 (22.6%) | 85,909 (40.6%) | 8,650 (5.3%) | 93,005 (63.5%) | 5,266 (4.2%) | 7,781 (9.3%) | 33,387 (40.3%) | 30,772 (40.2%) |
| Other | 355 (0.0%) | 21 (0.0%) | 0 (0%) | 21 (0.0%) | 0 (0%) | 1 (0.0%) | 52 (0.1%) | 33 (0.0%) |
| Rank |  |  |  |  |  |  |  |  |
| Enlisted | 1,477,717 (83.1%) | 187,474 (88.0%) | 134,912 (82.9%) | 120,546 (82.0%) | 108,030 (86.7%) | 70,755 (83.9%) | 65,936 (79.3%) | 66,053 (86.1%) |
| Officer | 275,678 (15.5%) | 24,013 (11.3%) | 24,716 (15.2%) | 24,401 (16.6%) | 14,634 (11.8%) | 12,206 (14.5%) | 16,458 (19.8%) | 9,590 (12.5%) |
| Warrant Officer | 25,968 (1.5%) | 1,641 (0.8%) | 3,036 (1.9%) | 1,987 (1.4%) | 1,908 (1.5%) | 1,419 (1.7%) | 747 (0.9%) | 1,040 (1.4%) |

Supplementary Table 3. Associations of demographics with the primary outcomes in the full study population

|  | Primary initiation timing | | Second primary dose timing | | Time between booster eligibility and receipt |
| --- | --- | --- | --- | --- | --- |
|  | Pre-mandate | Post-mandate | Recommended | Late | N=415,359 |
|  | N=1,230,623 (77.2%) | N=364,325 (22.8%) | N=1,389,085 (96.2%) | N=55,690 (3.9%) | 90 (68-120) |
| Age |  |  |  |  |  |
| 17-49 | **1,199,817 (76.9%)** | **360,566 (23.1%)** | 1,358,545 (96.2%) | 54,406 (3.9%) | **91 (68-120)** |
| 50-65 | **30,806 (89.1%)** | **3,759 (1.0%)** | 30,540 (96.0%) | 1,284 (4.0%) | **75 (54-98)** |
| Sex |  |  |  |  |  |
| Male | **1,007,738 (77.0%)** | **300,659 (23.0%)** | **1,131,477 (96.1%)** | **45,688 (3.9%)** | **91 (69-121)** |
| Female | **222,885 (77.8%)** | **63,666 (22.2%)** | **257,608 (96.3%)** | **10,002 (3.7%)** | **86 (61-114)** |
| Race |  |  |  |  |  |
| American Indian or Alaska Native | **12,819 (78.1%)** | **3,591 (21.9%)** | **14,325 (96.4%)** | **536 (3.6%)** | **98 (71-124)** |
| Asian or Pacific Islander | **87,816 (87.2%)** | **12,935 (12.8%)** | **89,357 (96.7%)** | **3,029 (3.3%)** | **90 (67-120)** |
| Black or African American | **187,887 (71.6%)** | **74,570 (28.4%)** | **230,537 (95.7%)** | **10,380 (4.3%)** | **91 (65-121)** |
| White | **855,764 (77.1%)** | **253,995 (22.9%)** | **960,602 (96.1%)** | **38,766 (3.9%)** | **90 (68-120)** |
| Other | **69,662 (81.9%)** | **15,399 (18.1%)** | **75,769 (96.9%)** | **2,451 (3.1%)** | **90 (67-118)** |
| Hispanic Ethnicity |  |  |  |  |  |
| Yes | **215,696 (77.5%)** | **62,684 (25.5%)** | 244,643 (96.1%) | 10,014 (3.9%) | **96 (69-125)** |
| No | **1,000,084 (77.1%)** | **297,955 (22.9%)** | 1,127,738 (96.2%) | 45,207 (3.9%) | **90 (67-119)** |
| Branch of Service |  |  |  |  |  |
| Air Force | **280,319 (75.0%)** | **93,523 (25.0%)** | **331,383 (96.5%)** | **12,203 (3.6%)** | **86 (67-113)** |
| Army | **474,566 (77.4%)** | **138,753 (22.6%)** | **522,054 (95.3%)** | **25,579 (4.7%)** | **87 (64-119)** |
| Coast Guard | **35,004 (84.0%)** | **6,648 (16.0%)** | **36,487 (95.6%)** | **1,667 (4.4%)** | **84 (58-111)** |
| Marine Corps | **130,560 (66.4%)** | **65,982 (33.6%)** | **171,502 (96.0%)** | **7,207 (4.0%)** | **106 (75-140)** |
| Navy | **303,044 (84.0%)** | **57,686 (16.0%)** | **319,677 (97.4%)** | **8,691 (2.7%)** | **98 (70-124)** |
| Other | **196 (73.7%)** | **70 (26.3%)** | **242 (94.2%)** | **15 (5.8%)** | **75 (61-104)** |
| Rank |  |  |  |  |  |
| Enlisted | **968,747 (74.0%)** | **340,874 (26.0%)** | **1,135,370 (95.9%)** | **48,014 (4.1%)** | **98 (70-125)** |
| Officer | **240,584 (92.4%)** | **19,760 (7.6%)** | **232,755 (97.2%)** | **6,824 (2.9%)** | **81 (61-107)** |
| Warrant Officer | **21,281 (85.2%)** | **3,689 (14.8%)** | **20,947 (96.1%)** | **852 (3.9%)** | **87 (68-113)** |
| COVID before primary initiation |  |  |  |  |  |
| Lab confirmed | **86,949 (67.0%)** | **42,842 (33.0%)** | **11,331 (85.5%)** | **1,925 (14.5%)** | **126 (98-169)** |
| ICD-10-CM defined | **79,270 (65.3%)** | **26,214 (7.2%)** | **8,800 (85.9%)** | **1,447 (14.1%)** | **107 (77-142)** |
| No | **1,094,395 (78.8%)** | **295,269 (21.3%)** | **1,368,954 (96.3%)** | **52,318 (3.7%)** | **87 (66-114)** |

^a^Bold text indicates a significant difference (P<0.05) across the comparison groups.

Supplementary Table 4. Demographics associated with primary initiation timing in relation to the DoD mandate in select states of residence^a^

|  | California | | Texas | | Virginia | | North Carolina | | Georgia | | Florida | | Washington | |
| --- | --- | --- | --- | --- | --- | --- | --- | --- | --- | --- | --- | --- | --- | --- |
|  | Pre-mandate | Post-mandate | Pre-mandate | Post-mandate | Pre-mandate | Post-mandate | Pre-mandate | Post-mandate | Pre-mandate | Post-mandate | Pre-mandate | Post-mandate | Pre-mandate | Post-mandate |
|  | N=147,271 (78.2%) | N=41,138 (21.8%) | N=108,818 (74.7%) | N=36,818 (25.3%) | N=108,256 (80.4%) | N=26,423 (19.6%) | N=75,147 (70.2%) | N=31,878 (29.8%) | N=55,405 (75.1%) | N=18,340 (24.9%) | N=56.734 (73.6%) | N=20,397 (26.4%) | N=58,248 (84.5%) | N=10,726 (15.5%) |
| Age |  |  |  |  |  |  |  |  |  |  |  |  |  |  |
| 17-49 | **145,391 (78.0%)** | **40,960 (22.0%)** | **105,826 (74.4%)** | **36,381 (25.6%)** | **105,449 (80.1%)** | **26,234 (19.9%)** | **74,000 (70.0%)** | **31,724 (30.0%)** | **54,159 (75.0%)** | **18,065 (25.0%)** | **55,392 (73.3%)** | **20,231 (26.8%)** | **57,500 (84.4%)** | **10,647 (15.6%)** |
| 50-65 | **1,880 (91.4%)** | **178 (8.7%)** | **2,992 (87.3%)** | **437 (12.7%)** | **2,807 (93.7%)** | **189 (6.3%)** | **1,147 (88.2%)** | **154 (11.8%)** | **1,246 (81.9%)** | **275 (18.1%)** | **1,342 (89.6%)** | **156 (10.4%)** | **748 (90.5%)** | **79 (9.6%)** |
| Sex |  |  |  |  |  |  |  |  |  |  |  |  |  |  |
| Male | **121,886 (77.7%)** | **34,896 (22.3%)** | 87,004 (74.7%) | 29,449 (25.3%) | **85,368 (80.6%)** | **20,568 (19.4%)** | **65,670 (69.8%)** | **28,467 (30.2%)** | **47,253 (75.4%)** | **15,428 (24.6%)** | 45,720 (73.5%) | 16,491 (26.5%) | **48,664 (84.7%)** | **8,773 (15.3%)** |
| Female | **25,385 (80.3%)** | **6,242 (19.7%)** | 21,814 (74.8%) | 7,369 (25.3%) | **22,888 (79.6%)** | **5,855 (20.4%)** | **9,477 (73.5%)** | **3,411 (26.5%)** | **8,152 (73.7%)** | **2,912 (26.3%)** | 11,014 (73.9%) | 3,896 (26.1%) | **9,584 (83.1%)** | **1,953 (16.9%)** |
| Race |  |  |  |  |  |  |  |  |  |  |  |  |  |  |
| American Indian or Alaska Native | **2,155 (79.3%)** | **564 (20.7%)** | **930 (74.9%)** | **311 (25.1%)** | **1,492 (81.3%)** | **344 (18.7%)** | **705 (71.7%)** | **279 (28.4%)** | **430 (79.5%)** | **111 (20.5%)** | **638 (76.2%)** | **199 (23.8%)** | **818 (84.8%)** | **147 (15.2%)** |
| Asian or Pacific Islander | **13,922 (88.0%)** | **1,901 (12.0%)** | **8,309 (85.7%)** | **1,389 (14.3%)** | **6,730 (88.4%)** | **885 (11.6%)** | **3,907 (81.0%)** | **917 (19.0%)** | **2,961 (84.4%)** | **549 (15.6%)** | **2,861 (83.6%)** | **560 (16.4%)** | **5,363 (90.4%)** | **572 (9.6%)** |
| Black or African American | **18,999 (74.1%)** | **6,629 (25.9%)** | **19,789 (69.6%)** | **8,657 (30.4%)** | **20,188 (74.5%)** | **6,923 (25.5%)** | **10,835 (67.5%)** | **5,217 (32.5%)** | **11,223 (68.5%)** | **5,157 (31.5%)** | **8,610 (66.1%)** | **4,419 (33.9%)** | **7,628 (78.7%)** | **2,062 (21.3%)** |
| White | **99,637 (77.2%)** | **29,472 (22.8%)** | **74,217 (75.2%)** | **24,486 (24.8%)** | **70,118 (81.3%)** | **16,168 (18.7%)** | **57,164 (70.0%)** | **24,499 (30.0%)** | **38,635 (76.7%)** | **11,760 (22.3%)** | **39,265 (74.1%)** | **13,745 (25.9%)** | **40,727 (84.8%)** | **7,322 (15.2%)** |
| Other | **11,691 (84.0%)** | **2,223 (16.0%)** | **4,019 (75.7%)** | **1,289 (24.3%)** | **8,856 (82.5%)** | **1,881 (17.5%)** | **2,024 (73.7%)** | **722 (26.3%)** | **1,598 (76.0%)** | **506 (24.1%)** | **4,574 (78.9%)** | **1,224 (21.1%)** | **3,428 (85.9%)** | **561 (14.1%)** |
| Hispanic Ethnicity |  |  |  |  |  |  |  |  |  |  |  |  |  |  |
| Yes | **35,777 (76.3%)** | **11,120 (23.7%)** | **24,340 (76.2%)** | **7,613 (23.8%)** | 16,754 (80.5%) | 4,050 (19.5%) | **13,185 (69.5%)** | **5,783 (30.5%)** | **7,857 (76.8%)** | **2,379 (23.2%)** | 10,025 (73.8%) | 3,565 (26.2%) | **10,056 (85.2%)** | **1,743 (14.8%)** |
| No | **110,711 (78.9%)** | **29,678 (21.1%)** | **83,157 (74.5%)** | **28,543 (25.6%)** | 90,726 (80.4%) | 22,161 (19.6%) | **61,590 (70.4%)** | **25,863 (29.6%)** | **47,137 (75.0%)** | **15,716 (25.0%)** | 45,993 (73.5%) | 16,597 (26.5%) | **47,978 (84.3%)** | **8,926 (15.7%)** |
| Branch of Service |  |  |  |  |  |  |  |  |  |  |  |  |  |  |
| Air Force | **16,660 (77.2%)** | **4,912 (22.8%)** | **26,772 (68.9%)** | **12,078 (31.1%)** | **7,706 (79.2%)** | **2,022 (20.8%)** | **4,347 (68.9%)** | **1,962 (31.1%)** | **7,581 (73.2%)** | **2,781 (26.8%)** | **17,183 (68.0%)** | **8,093 (32.0%)** | **5,657 (70.4%)** | **2,376 (29.6%)** |
| Army | **11,512 (76.9%)** | **3,452 (23.1%)** | **72,016 (77.2%)** | **21,273 (22.8%)** | **17,251 (76.7%)** | **5,254 (23.4%)** | **37,683 (82.0%)** | **8,275 (18.0%)** | **40,278 (74.9%)** | **13,496 (25.1%)** | **7,536 (69.5%)** | **3,306 (30.5%)** | **26,045 (84.7%)** | **4,716 (15.3%)** |
| Coast Guard | **4,305 (87.7%)** | **602 (12.3%)** | **1,366 (78.0%)** | **386 (22.0%)** | **3,764 (86.4%)** | **593 (13.6%)** | **1,197 (76.6%)** | **365 (23.4%)** | **380 (78.5%)** | **104 (21.5%)** | **4,007 (80.7%)** | **959 (19.3%)** | **1,447 (87.4%)** | **209 (12.6%)** |
| Marine Corps | **45,761 (68.4%)** | **21,133 (31.6%)** | **2,300 (69.9%)** | **990 (30.1%)** | **8,806 (72.3%)** | **3,382 (27.8%)** | **28,041 (58.1%)** | **20,253 (41.9%)** | **1,048 (67.8%)** | **497 (32.2%)** | **3,135 (67.5%)** | **1,510 (32.5%)** | **706 (76.0%)** | **223 (24.0%)** |
| Navy | **67,981 (86.2%)** | **10,880 (13.8%)** | **6,024 (75.7%)** | **1,938 (24.3%)** | **70,285 (82.3%)** | **15,125 (17.7%)** | **3,846 (79.6%)** | **983 (3.1%)** | **6,039 (83.4%)** | **1,203 (16.6%)** | **24,631 (79.2%)** | **6,476 (20.8%)** | **24,311 (88.4%)** | **3,182 (11.6%)** |
| Other | **11 (61.1%)** | **7 (38.9%)** | **0 (0%)** | **0 (0%)** | **7 (50.0%)** | **7 (50.0%)** | **-** | **-** | **1 (100%)** | **0 (0%)** | **38 (82.6%)** | **8 (17.4%)** | **20 (76.9%)** | **6 (23.1%)** |
| Rank |  |  |  |  |  |  |  |  |  |  |  |  |  |  |
| Enlisted | **124,149 (75.8%)** | **39,660 (24.2%)** | **85,119 (71.4%)** | **34,120 (28.6%)** | **84,764 (77.4%)** | **24,770 (22.6%)** | **60,993 (66.7%)** | **30,447 (33.3%)** | **44,487 (72.5%)** | **16,890 (27.5%)** | **41,637 (68.8%)** | **18,880 (31.2%)** | **48,675 (82.7%)** | **10,165 (17.3%)** |
| Officer | **21,690 (94.3%)** | **1,303 (5.7%)** | **21,298 (90.8%)** | **2,167 (9.2%)** | **21,795 (93.8%)** | **1,446 (6.2%)** | **12,621 (91.8%)** | **1,128 (8.2%)** | **9,766 (88.8%)** | **1,232 (11.2%)** | **14,487 (91.2%)** | **1,407 (8.9%)** | **8,661 (94.9%)** | **465 (5.1%)** |
| Warrant Officer | **1,432 (89.1%)** | **175 (10.9%)** | **2,398 (81.9%)** | **530 (18.1%)** | **1,697 (89.2%)** | **206 (10.8%)** | **1,532 (83.5%)** | **303 (16.5%)** | **1,152 (84.1%)** | **218 (15.9%)** | **609 (85.9%)** | **100 (14.1%)** | **912 (90.5%)** | **96 (9.5%)** |
| COVID before primary initiation |  |  |  |  |  |  |  |  |  |  |  |  |  |  |
| Lab confirmed | **9,665 (68.5%)** | **4,438 (31.5%)** | **9,315 (68.4%)** | **4,307 (31.6%)** | **7,710 (72.3%)** | **2,954 (27.7%)** | **6,321 (56.9%)** | **4,784 (43.1%)** | **5,869 (65.1%)** | **3,149 (34.9%)** | **4,587 (61.4%)** | **2,886 (38.6%)** | **3,317 (72.2%)** | **1,280 (27.8%)** |
| ICD-10-CM defined | **3,958 (68.7%)** | **1,807 (31.3%)** | **6,100 (69.0%)** | **2,744 (31.0%)** | **3,166 (65.2%)** | **1,689 (34.8%)** | **3,006 (64.9%)** | **1,626 (35.1%)** | **2,142 (66.3%)** | **1,088 (33.7%)** | **2,254 (52.3%)** | **2,054 (47.7%)** | **1,383 (70.6%)** | **577 (29.4%)** |
| No | **133,648 (68.7%)** | **34.893 (20.7%)** | **93,403 (75.8%)** | **29,767 (24.2%)** | **97,380 (81.7%)** | **21,780 (18.3%)** | **65,820 (72.1%)** | **25,468 (27.9%)** | **47,394 (77.1%)** | **14,103 (22.9%)** | **49,893 (76.4%)** | **15,447 (23.6%)** | **53,548 (85.8%)** | **8,869 (14.2%)** |

^a^Bold text indicates a significant difference (P<0.05) across the comparison groups within the indicated state.

Supplementary Table 5. Unadjusted odds of completing the second dose of a two-dose primary series in the manufacturer-recommended timeframe, overall and in select states of residence^a,b^

|  | All | California | Texas | Virginia | North Carolina | Georgia | Florida | Washington |
| --- | --- | --- | --- | --- | --- | --- | --- | --- |
|  | N=1,444,775 | N=171,978 | N=131,004 | N=124,257 | N=98,234 | N=66,499 | N=70,350 | N=60,715 |
|  | Univariate  OR (95% CI) | Univariate  OR (95% CI) | Univariate  OR (95% CI) | Univariate  OR (95% CI) | Univariate  OR (95% CI) | Univariate  OR (95% CI) | Univariate  OR (95% CI) | Univariate  OR (95% CI) |
| Age |  |  |  |  |  |  |  |  |
| 17-49 | Ref | Ref | Ref | Ref | Ref | Ref | Ref | Ref |
| 50-65 | 0.95 (0.90-1.01) | **0.71 (0.58-0.87)** | 1.05 (0.89-1.24) | **1.76 (1.30-2.37)** | **1.47 (1.04-2.08)** | **0.74 (0.59-0.94)** | 0.99 (0.72-1.36) | 0.91 (0.60-1.37) |
| Sex |  |  |  |  |  |  |  |  |
| Male | Ref | Ref | Ref | Ref | Ref | Ref | Ref | Ref |
| Female | **1.04 (1.02-1.06)** | **1.16 (1.09-1.24)** | **1.12 (1.05-1.19)** | 0.97 (0.89-1.05) | 0.98 (0.89-1.08) | 0.99 (0.89-1.10) | 0.95 (0.86-1.07) | 0.92 (0.82-1.04) |
| Race |  |  |  |  |  |  |  |  |
| American Indian or Alaska Native | 1.08 (0.99-1.18) | **1.35 (1.07-1.70)** | 0.89 (0.69-1.15) | 1.36 (0.96-1.94) | 0.75 (0.56-1.01) | 0.92 (0.59-1.44) | 1.09 (0.70-1.70) | 0.72 (0.51-1.02) |
| Asian or Pacific Islander | **1.19 (1.15-1.24)** | 1.02 (0.94-1.12) | **1.23 (1.11-1.38)** | **1.65 (1.36-2.00)** | **1.18 (1.00-1.39)** | 1.16 (0.95-1.41) | 1.26 (0.99-1.60) | 1.11 (0.94-1.33) |
| Black or African American | **0.90 (0.88-0.92)** | 1.01 (0.94-1.09) | 0.95 (0.89-1.01) | **0.74 (0.68-0.80)** | **0.90 (0.82-0.98)** | **0.77 (0.70-0.84)** | **0.88 (0.79-0.99)** | **0.83 (0.73-0.94)** |
| White | Ref | Ref | Ref | Ref | Ref | Ref | Ref | Ref |
| Other | **1.25 (1.20-1.30)** | **1.23 (1.11-1.36)** | **1.40 (1.20-1.63)** | 0.99 (0.87-1.13) | **1.28 (1.02-1.61)** | 0.96 (0.76-1.22) | **1.37 (1.13-1.67)** | 1.16 (0.94-1.44) |
| Hispanic Ethnicity |  |  |  |  |  |  |  |  |
| Yes | 0.98 (0.96-1.00) | **0.92 (0.87-0.97)** | 1.02 (0.96-1.08) | **0.90 (0.82-0.99)** | 0.95 (0.88-1.03) | 1.07 (0.95-1.20) | 0.90 (0.80-1.00) | 1.05 (0.93-1.19) |
| No | Ref | Ref | Ref | Ref | Ref | Ref | Ref | Ref |
| Branch of Service |  |  |  |  |  |  |  |  |
| Air Force | **1.33 (1.30-1.36)** | **2.35 (2.12-2.60)** | **1.58 (1.49-1.68)** | 0.96 (0.83-1.10) | **0.83 (0.72-0.94)** | 0.97 (0.87-1.08) | **1.48 (1.32-1.67)** | **1.19 (1.02-1.39)** |
| Army | Ref | Ref | Ref | Ref | Ref | Ref | Ref | Ref |
| Coast Guard | **1.07 (1.02-1.13)** | **1.61 (1.38-1.88)** | **1.38 (1.08-1.77)** | 1.11 (0.90-1.35) | **1.96 (1.35-2.86)** | **2.11 (1.09-4.09)** | **2.17 (1.74-2.70)** | **0.31 (0.26-0.37)** |
| Marine Corps | **1.17 (1.14-1.20)** | **1.60 (1.48-1.73)** | **1.74 (1.42-2.12)** | **1.16 (1.01-1.34)** | 0.94 (0.88-1.00) | **1.39 (1.03-1.89)** | **1.52 (1.25-1.85)** | 1.04 (0.71-1.52) |
| Navy | **1.80 (1.76-1.85)** | **2.38 (2.20-2.57)** | **1.97 (1.71-2.26)** | **1.29 (1.18-1.41)** | 1.16 (0.98-1.37) | **2.44 (2.03-2.93)** | **2.28 (2.01-2.59)** | **1.73 (1.55-1.93)** |
| Rank |  |  |  |  |  |  |  |  |
| Enlisted | Ref | Ref | Ref | Ref | Ref | Ref | Ref | Ref |
| Officer | **1.44 (1.41-1.48)** | 1.03 (0.95-1.11) | **1.61 (1.49-1.74)** | **2.01 (1.79-2.26)** | **1.75 (1.56-1.97)** | **1.34 (1.19-1.51)** | **1.69 (1.49-1.92)** | 1.01 (0.88-1.16) |
| Warrant Officer | 1.04 (0.97-1.11) | 0.92 (0.72-1.19) | 1.15 (0.95-1.39) | **2.35 (1.54-3.58)** | **1.56 (1.16-2.11)** | **1.40 (1.01-1.95)** | 1.26 (0.77-2.08) | **0.50 (0.38-0.66)** |
| COVID infection between primary doses |  |  |  |  |  |  |  |  |
| Lab confirmed | **0.23 (0.21-0.24)** | **0.21 (0.18-0.24)** | **0.26 (0.23-0.30)** | **0.24 (0.18-0.31)** | **0.18 (0.15-0.21)** | **0.39 (0.32-0.48)** | **0.14 (0.11-0.18)** | **0.25 (0.19-0.33)** |
| ICD-10-CM defined | **0.23 (0.22-0.25)** | **0.26 (0.21-0.32)** | **0.33 (0.28-0.40)** | **0.19 (0.15-0.25)** | **0.21 (0.17-0.28)** | **0.23 (0.17-0.29)** | **0.13 (0.10-0.17)** | **0.24 (0.17-0.35)** |
| No | Ref | Ref | Ref | Ref | Ref | Ref | Ref | Ref |

^a^Bold text indicates a significant difference from the reference category at P<0.05. ^b^Those who received a one-dose Johnson & Johnson/Janssen primary vaccine dose were excluded from these analyses

OR, odds ratio; CI, confidence interval; Ref, reference group.

Supplementary Table 6. Demographics associated with second primary dose timing in relation to manufacturer recommendations in select states of residence^a^

|  | California | | Texas | | Virginia | | North Carolina | | Georgia | | Florida | | Washington | |
| --- | --- | --- | --- | --- | --- | --- | --- | --- | --- | --- | --- | --- | --- | --- |
|  | Recommended | Late | Recommended | Late | Recommended | Late | Recommended | Late | Recommended | Late | Recommended | Late | Recommended | Late |
|  | N=165,245 (96.1%) | N=6,733 (3.9%) | N=124,558 (95.1%) | N=6,446 (4.9%) | N=120,944 (97.3%) | N=3,313 (2.7%) | N=94,342 (96.0%) | N=3,892 (4.0%) | N=63,816 (96.0%) | N=2,683 (4.0%) | N=68,325 (97.1%) | N=2,025 (2.9%) | N=60,715 (97.0%) | N=1,862 (3.0%) |
| Age |  |  |  |  |  |  |  |  |  |  |  |  |  |  |
| 17-49 | **163,404 (96.1%)** | **6,628 (3.9%)** | 121,643 (95.1%) | 6,302 (4.9%) | **118,153 (97.3%)** | **3,269 (2.7%)** | **93,172 (96.0%)** | **3,859 (4.0%)** | **62,496 (96.0%)** | **2,609 (4.0%)** | 66,985 (97.1%) | 1,985 (2.9%) | 60,004 (97.0%) | 1,838 (3.0%) |
| 50-65 | **1,841 (94.6%)** | **105 (5.4%)** | 2,915 (95.3%) | 144 (4.7%) | **2,791 (98.5%)** | **44 (1.6%)** | **1,170 (97.3%)** | **33 (2.7%)** | **1,320 (95.7%)** | **74 (5.3%)** | 1,340 (97.1%) | 40 (2.9%) | 711 (96.7%) | 24 (3.3%) |
| Sex |  |  |  |  |  |  |  |  |  |  |  |  |  |  |
| Male | **136,541 (96.0%)** | **5,701 (4.0%)** | **98,699 (95.0%)** | **5,221 (5.0%)** | 94,377 (97.4%) | 2,565 (2.7%) | 82,642 (96.0%) | 3,399 (4.0%) | 53,953 (96.0%) | 2,265 (4.0%) | 54,737 (97.2%) | 1,607 (2.9%) | 50,407 (97.1%) | 1,523 (2.9%) |
| Female | **28,704 (96.5%)** | **1,032 (3.5%)** | **25,859 (95.5%)** | **1,225 (4.5%)** | 36,567 (97.3%) | 748 (2.7%) | 11,700 (96.0%) | 493 (4.0%) | 9,863 (95.9%) | 418 (4.1%) | 13,588 (97.0%) | 418 (3.0%) | 10,308 (96.8%) | 339 (3.2%) |
| Race |  |  |  |  |  |  |  |  |  |  |  |  |  |  |
| American Indian or Alaska Native | **2,420 (97.0%)** | **75 (3.0%)** | **1,052 (94.4%)** | **62 (5.6%)** | **1,662 (98.1%)** | **32 (1.9%)** | **845 (94.8%)** | **46 (5.2%)** | **464 (95.9%)** | **20 (4.1%)** | **731 (97.3%)** | **20 (2.7%)** | **839 (96.0%)** | **35 (4.0%)** |
| Asian or Pacific Islander | **14,113 (96.1%)** | **575 (3.9%)** | **8,494 (95.9%)** | **362 (4.1%)** | **7,028 (98.4%)** | **112 (1.6%)** | **4,359 (96.7%)** | **151 (3.4%)** | **3,090 (96.7%)** | **106 (3.3%)** | **3,096 (97.7%)** | **73 (2.3%)** | **5,353 (97.4%)** | **145 (2.6%)** |
| Black or African American | **22,740 (96.1%)** | **936 (4.0%)** | **24,593 (94.8%)** | **1,364 (5.3%)** | **24,565 (96.6%)** | **874 (3.4%)** | **14,385 (95.6%)** | **657 (4.4%)** | **14,311 (95.1%)** | **741 (4.9%)** | **11,796 (96.7%)** | **397 (3.3%)** | **8,506 (96.5%)** | **309 (3.5%)** |
| White | **112,533 (96.0%)** | **4,696 (4.0%)** | **83,680 (95.0%)** | **4,397 (5.0%)** | **77,046 (97.4%)** | **2,024 (2.6%)** | **71,633 (96.1%)** | **2,930 (3.9%)** | **43,381 (96.2%)** | **1,719 (3.8%)** | **46,549 (97.1%)** | **1,385 (2.9%)** | **42,251 (97.1%)** | **1,275 (2.9%)** |
| Other | **12,380 (96.7%)** | **421 (3.3%)** | **4,752 (96.4%)** | **178 (3.6%)** | **9,689 (97.4%)** | **256 (2.6%)** | **2,449 (96.9%)** | **78 (3.1%)** | **1,848 (96.1%)** | **76 (4.0%)** | **5,267 (97.9%)** | **114 (2.1%)** | **3,466 (97.5%)** | **90 (2.5%)** |
| Hispanic Ethnicity |  |  |  |  |  |  |  |  |  |  |  |  |  |  |
| Yes | **41,460 (95.8%)** | **1,807 (4.2%)** | 27,626 (95.1%) | 1,417 (4.9%) | **18,814 (97.1%)** | **564 (2.9%)** | 16,875 (95.9%) | 725 (4.1%) | 8,901 (96.2%) | 355 (3.8%) | 12,180 (96.9%) | 393 (3.1%) | 10,434 (97.1%) | 308 (2.9%) |
| No | **122,808 (96.2%)** | **4,903 (4.2%)** | 95,173 (95.1%) | 4,954 (5.0%) | **101,272 (97.4%)** | **2,735 (2.6%)** | 76,937 (96.1%) | 3,141 (3.9%) | 54,338 (95.9%) | 2,310 (4.1%) | 55,337 (97.2%) | 1,598 (2.8%) | 50,047 (97.0%) | 1,550 (3.0%) |
| Branch of Service |  |  |  |  |  |  |  |  |  |  |  |  |  |  |
| Air Force | **19,380 (96.9%)** | **620 (3.1%)** | **34,838 (96.3%)** | **1,331 (3.7%)** | **8,866 (96.7%)** | **301 (3.3%)** | **5,557 (95.4%)** | **270 (4.6%)** | **8,992 (95.6%)** | **414 (4.4%)** | **22,354 (96.8%)** | **732 (3.2%)** | **7,106 (97.1%)** | **210 (2.9%)** |
| Army | **12,919 (93.0%)** | **969 (7.0%)** | **77,850 (94.3%)** | **4,705 (5.7%)** | **20,075 (96.9%)** | **651 (3.1%)** | **40,491 (96.1%)** | **1,624 (3.9%)** | **46,111 (95.7%)** | **2,056 (4.3%)** | **9,156 (95.4%)** | **445 (4.6%)** | **27,731 (96.6%)** | **976 (3.4%)** |
| Coast Guard | **4,408 (95.6%)** | **205 (4.4%)** | **1,505 (95.8%)** | **66 (4.2%)** | **3,920 (97.2%)** | **115 (2.9%)** | **1,370 (98.0%)** | **28 (2.0%)** | **426 (97.9%)** | **9 (2.1%)** | **4,409 (97.8%)** | **99 (2.2%)** | **1,377 (89.8%)** | **156 (10.2%)** |
| Marine Corps | **57,989 (95.5%)** | **2,717 (4.5%)** | **2,842 (96.6%)** | **99 (3.4%)** | **10,864 (97.3%)** | **303 (2.7%)** | **42,466 (95.9%)** | **1,816 (4.1%)** | **1,376 (96.9%)** | **44 (3.1%)** | **4,098 (96.9%)** | **131 (3.1%)** | **825 (96.7%)** | **28 (3.3%)** |
| Navy | **69,444 (46.9%)** | **2,190 (3.1%)** | **7,093 (97.0%)** | **218 (3.0%)** | **76,758 (97.5%)** | **1,935 (2.5%)** | **4,392 (96.7%)** | **152 (3.4%)** | **6,615 (98.2%)** | **121 (1.8%)** | **28,066 (97.9%)** | **598 (2.1%)** | **23,586 (98.0%)** | **481 (2.0%)** |
| Other | **18 (100%)** | **0 (0%)** | **0 (0%)** | **0 (0%)** | **13 (100%)** | **0 (0%)** | **-** | **-** | **1 (100%)** | **0 (0%)** | **37 (80.4%)** | **9 (19.6%)** | **25 (95.2%)** | **1 (3.9%)** |
| Rank |  |  |  |  |  |  |  |  |  |  |  |  |  |  |
| Enlisted | 143,323 (96.1%) | 5,855 (3.9%) | **101,509 (94.8%)** | **5,621 (5.3%)** | **97,876 (97.1%)** | **2,969 (2.9%)** | **80,353 (95.8%)** | **3,536 (4.2%)** | **53,025 (95.8%)** | **2,331 (4.2%)** | **53,401 (96.9%)** | **1,734 (3.2%)** | **51,589 (97.1%)** | **1,562 (2.9%)** |
| Officer | 20,521 (96.2%) | 816 (3.8%) | **20,673 (96.7%)** | **711 (3.3%)** | **21,365 (98.5%)** | **322 (1.5%)** | **12,390 (97.6%)** | **311 (2.5%)** | **9,609 (96.8%)** | **315 (3.2%)** | **14,301 (98.1%)** | **275 (1.9%)** | **8,249 (97.1%)** | **247 (2.9%)** |
| Warrant Officer | 1,401 (95.8%) | 62 (4.2%) | **2,372 (95.4%)** | **114 (4.6%)** | **1,702 (98.7%)** | **22 (1.3%)** | **1,598 (97.3%)** | **45 (2.7%)** | **1,182 (97.0%)** | **37 (3.0%)** | **622 (97.5%)** | **16 (2.5%)** | **877 (94.3%)** | **53 (5.7%)** |
| COVID between first and second primary dose |  |  |  |  |  |  |  |  |  |  |  |  |  |  |
| Lab confirmed | **1,177 (84.2%)** | **221 (15.8%)** | **1,227 (83.9%)** | **235 (16.1%)** | **558 (90.0%)** | **62 (10.0%)** | **785 (82.2%)** | **170 (17.8%)** | **1,125 (90.7%)** | **115 (9.3%)** | **366 (83.6%)** | **72 (16.4%)** | **551 (89.5%)** | **65 (10.6%)** |
| ICD-10-CM defined | **592 (86.7%)** | **91 (13.3%)** | **985 (87.1%)** | **146 (12.9%)** | **483 (88.0%)** | **66 (12.0%)** | **393 (84.5%)** | **72 (15.5%)** | **382 (84.9%)** | **68 (15.1%)** | **351 (82.2%)** | **76 (17.8%)** | **225 (89.2%)** | **31 (10.8%)** |
| No | **163,476 (96.2%)** | **6,421 (3.8%)** | **122,346 (95.3%)** | **6,065 (4.7%)** | **119,903 (97.4%)** | **3,185 (2.6%)** | **93,164 (96.2%)** | **3,650 (3.8%)** | **62,309 (96.1%)** | **2,500 (3.9%)** | **67,608 (97.3%)** | **1,877 (2.7%)** | **59,909 (97.1%)** | **1,766 (2.9%)** |

^a^Bold text indicates a significant difference (P<0.05) across the comparison groups within the indicated state.

Supplementary Table 7. Demographics associated with time between booster eligibility and receipt in select states of residence^a^

|  | California | Texas | Virginia | North Carolina | Georgia | Florida | Washington |
| --- | --- | --- | --- | --- | --- | --- | --- |
|  | 98 (71-125) | 85 (99-115) | 88 (63-118) | 94 (63-127) | 96 (70-125) | 95 (70-120) | 98 (72-121) |
| Age |  |  |  |  |  |  |  |
| 17-49 | **99 (71-125)** | **86 (60-116)** | **89 (65-120)** | **96 (65-127)** | **97 (71-126)** | **96 (70-121)** | **98 (73-122)** |
| 50-65 | **76 (52-105)** | **73 (49-100)** | **72 (49-92)** | **76 (51-104)** | **82 (62-106)** | **79 (58-104)** | **77 (51-105)** |
| Sex |  |  |  |  |  |  |  |
| Male | **99 (71-125)** | **86 (61-117)** | **89 (65-120)** | **96 (65-128)** | **97 (71-126)** | **96 (71-120)** | **98 (73-122)** |
| Female | **96 (66-124)** | **81 (54-111)** | **84 (57-114)** | **89 (58-120)** | **90 (68-120)** | **91 (65-118)** | **96 (64-121)** |
| Race |  |  |  |  |  |  |  |
| American Indian or Alaska Native | **105 (76-125)** | **88 (63-125)** | 96 (65-127) | 97 (57-126) | **110 (74-135)** | 102 (69-125) | **107 (78-127)** |
| Asian or Pacific Islander | **97 (69-124)** | **84 (60-114)** | 87 (65-115) | 97 (63-126) | **95 (71-121)** | 95 (68-121) | **98 (72-121)** |
| Black or African American | **100 (71-125)** | **85 (58-114)** | 89 (60-120) | 97 (63-126) | **92 (68-120)** | 97 (69-121) | **105 (74-125)** |
| White | **98 (71-125)** | **86 (60-117)** | 87 (64-118) | 93 (63-127) | 97 (71-126) | 94 (70-120) | **97 (72-121)** |
| Other | **99 (71-125)** | **81 (56-107)** | 90 (66-119) | 92 (64-125) | 99 (71-131) | 95 (71-121) | **104 (75-125)** |
| Hispanic Ethnicity |  |  |  |  |  |  |  |
| Yes | **103 (72-125)** | **89 (63-119)** | **92 (65-123)** | **99 (69-132)** | **97 (72-132)** | 96 (69-122) | **103 (76-127)** |
| No | **98 (70-124)** | **84 (58-114)** | **87 (63-118)** | **92 (63-126)** | **96 (70-124)** | 95 (70-120) | **98 (72-121)** |
| Branch of Service |  |  |  |  |  |  |  |
| Air Force | **86 (61-113)** | **79 (54-107)** | **82 (62-106)** | **89 (69-118)** | **95 (72-120)** | **91 (71-116)** | **89 (72-118)** |
| Army | **75 (58-105)** | **89 (61-119)** | **79 (57-110)** | **95 (62-124)** | **91 (68-122)** | **85 (65-111)** | **97 (66-119)** |
| Coast Guard | **83 (56-114)** | **89 (58-115)** | **85 (58-111)** | **83 (64-110)** | **89 (62-109)** | **96 (76-119)** | **71 (43-104)** |
| Marine Corps | **106 (75-138)** | **91 (70-112)** | **99 (73-152)** | **99 (66-138)** | **98 (79-124)** | **90 (68-118)** | **106 (78-133)** |
| Navy | **105 (78-125)** | **87 (66-112)** | **92 (65-122)** | **90 (59-120)** | **108 (81-147)** | **98 (70-124)** | **105 (78-126)** |
| Other | **65 (41-142)** | **-** | **86 (71-126)** | **-** | **68 (-)** | **91 (71-125)** | **81 (72-118)** |
| Rank |  |  |  |  |  |  |  |
| Enlisted | **105 (74-126)** | **90 (62-120)** | **97 (67-129)** | **102 (66-133)** | **100 (71-131)** | **100 (71-127)** | **104 (76-126)** |
| Officer | **85 (63-110)** | **78 (54-105)** | **78 (58-104)** | **85 (61-113)** | **87 (69-113)** | **87 (68-111)** | **85 (59-111)** |
| Warrant Officer | **92 (69-117)** | **85 (62-112)** | **84 (62-110)** | **90 (68-114)** | **91 (75-114)** | **92 (71-113)** | **97 (69-116)** |
| COVID infection variable |  |  |  |  |  |  |  |
| Lab confirmed | **125 (99-167)** | **120 (85-160)** | **132 (100-189)** | **129 (97-168)** | **131 (101-174)** | **127 (105-193)** | **124 (98-160)** |
| ICD-10-CM defined | **112 (79-147)** | **96 (68-127)** | **113 (84-156)** | **111 (75-145)** | **110 (81-156)** | **116 (84-163)** | **113 (85-140)** |
| No | **97 (70-124)** | **83 (57-111)** | **86 (62-114)** | **90 (62-120)** | **91 (69-120)** | **91 (68-115)** | **96 (71-119)** |

^a^Bold text indicates a significant difference (P<0.05) across the comparison groups within the indicated state.
